# Trend analysis, modelling and impact assessment of COVID-19 in Nepal

**DOI:** 10.1101/2020.05.29.20117390

**Authors:** Shital Bhandary, Srijan Lal Shrestha, Ram Prasad Khatiwada, Deep Narayan Shah, Nabin Narayan Munankarmi, Megha Raj Banjara, Resham Thapa-Parajuli, Krishna Das Manandhar, Rameshwar Adhikari, Reshma Tuladhar

## Abstract

With continued global expansion of COVID-19 transmission and mounting threat of the the timely analysis of its trend in Nepal and forecasting the potential situation in the country has been deemed necessary. We analyzed the trend, modelling and impact assessment of COVID-19 disease, cases of Nepal from 23^rd^ January 2020 to 30^th^ April 2020 to portray the scenario of COVID-19 after the first phase of lockdown. Exponential smoothing state-space and autoregressive integrated moving average (ARIMA) models were constructed to forecast the cases. Susceptible-infectious-recovered (SIR) model was fit to estimate the basic reproduction number (Ro) of COVID-19 in Nepal. There has been increase in the number of cases but the overall growth in COVID-19 was not high. Statistical modelling has shown that COVID-19 cases may continue to increase exponentially in Nepal. The basic reproduction number in Nepal being maintained at low level of 1.08 for the period of 23^rd^ January to 30^th^ April 2020 is an indication of effectiveness of lockdown in containing the COVID-19 spread. The models further suggest that COVID-19 might persist until December 2020 with peak cases in August 2020. On the other hand, basic reproduction number of 1.25 was computed for total cases reported for the 22^nd^ March to 30^th^ April 2020 period implying that COVID-19 may remain for at least for a year in the country. Thus, maintaining social distance and stay home policy with an implementation of strict lockdown in COVID-19 affected district is highly recommended.

## Introduction

The ongoing Corona Virus Disease 2019 (COVID-19) caused by novel Severe Acute Respiratory Syndrome Coronavirus 2 (SARS CoV-2) was reported to have emerged from Wuhan, Hubei, China in late December 2019, where people in a seafood-wholesale wet market suffered from a mysterious pneumonia [1]. World Health Organization (WHO) declared the outbreak as public health emergency of international concern on 30^th^ January 2020, announced a new name for this disease as COVID-19 on 11^th^ February 2020 and pandemic on 11^th^ March 2020 [2].

With rapid spread in Europe, America and Asia, COVID-19 was confirmed globally in 3,090,445 people with 217,769 deaths by 30^th^ April 2020 [3]. The highest infection and death toll have been recorded in the USA followed by countries in Europe, Eastern Mediterranean, South-East Asia, Western Pacific and least in Africa [4].

The first case of COVID-19 in Nepal reported in a 32 years old Nepalese male who was admitted to hospital upon exhibiting mild symptoms, later discharged on 17^th^ January 2020 after improvement in clinical condition was a returnee from Wuhan City on 9^th^ January 2020 [5]. On 24^th^ January 2020 the infection was officially confirmed COVID-19 after reported positive from WHO reference laboratory in Hong Kong [5, 6]. Later the second case was confirmed on 23^rd^ March 2020 in a 19 years old female who returned from France via Doha, Qatar [7]. On 24^th^ March 2020 the Government of Nepal implemented nationwide lockdown advising residents to stay at home.

Until 13^th^ April 2020, the cases in Nepal were reported from the people who recently returned from abroad, and Indian nationals residing in Nepal where the latter case was in majority. The first case of indigenous transmission confirmed on 4^th^ April 2020 in a 34-year-old woman from Kailali District was a relative of the infected patient travelled from India [7]. By the end of the first phase of lock down of 28^th^ April 2020, 54 cases have been confirmed with documentation of no deaths [6]. COVID-19 has spread in 10 Districts of Nepal with Udayapur reported the maximum number of 25 cases.

Considered as the standard laboratory test for the diagnosis of COVID-19 the cases were confirmed by Reverse Transcription Real Time Polymerase Chain Reaction (RT-PCR) [8, 9]. Until 30^th^ April 2020, a total of 12,011 RT-PCR tests and 46860 Rapid Diagnostic Test (RDT) were performed in Nepal out of which 57 were tested positive. In consideration of numerous COVID-19 cases being detected in Nepal it has been essential to let the Nepal Government be prepared for the most predictable upcoming situation to tackle the COVID-19 pandemic. This paper, therefore, assessed the descriptive and trend analysis on growth of cases, its doubling time, statistical and epidemiological model and impacts.

## Materials and Methods

### Data sources

The secondary data from Ministry of Health and Population (MoHP), Nepal, Health Emergency Operation Centre, and data from relevant websites related to COVID-19 in Nepal were analyzed. Trend analysis was performed from daily compiled data of 102 days from the last week of January 2020 till the end of April 2020. Major variables associated with the disease; such as demographic variables of the cases, diagnostic tests including RT-PCR and RDT, number of cumulative and daily cases as well as that of recovered, quarantined and isolated cases were considered. Additionally, analysis of important variables such as cases as percent of PCR tests, doubling time of COVID-19 cases for Nepal was weekly assessed and represented graphically. Finally, different time points were used to develop predication models while impact assessments have been analyzed using the whole data.

### Statistical analysis: ETS and ARIMA models

The exponential smoothing state-space method (which basically comprises ‘Error, Trend, and Seasonal’ components in smoothing procedure of an event under consideration; hence named ETS model) which takes information into comprehensive consideration and ARIMA model [10] were used to forecast COVID-19 cases for Nepal by fitting in R software using forecast package [11]. We selected the best ETS model and best ARIMA model for 99 days official cumulative COVID-19 cases from 23^rd^ January to 30^th^ April 2020 and forecasted the cases for next 14 days (1^st^ May – 14^th^ May 2020). An attempt has been made in the direction of epidemiological modelling through application of SIR model (see next section for detail) based upon the suitable parameter values for Nepal.

### Mathematical analysis: SIR model

We used SIR model, a compartmental model where population is divided into three compartments: susceptible, infectious and recovered. We need two types of rates to move from one compartment to other viz. rate of transmission (β) to move from Susceptible to Infectious compartment and rate of recovery (γ) to move Infectious to Recovered compartment as follows

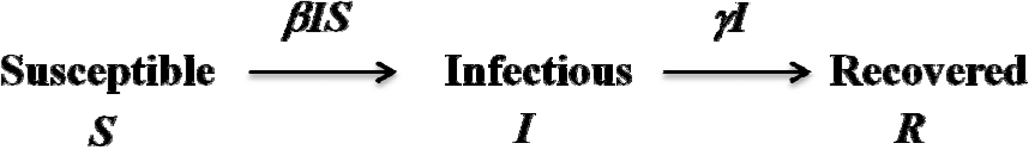

This type of model is a dynamic model as the population in these three compartments changes with each unit of time and this can be modelled using differential equations. The simple SIR model without vital dynamics i.e. births and deaths are modelled as follows:

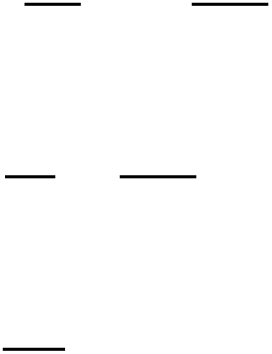

Where, S, I, and R were stock of susceptible, infected and recovered population. Change in S, I and R with respect to time was computed with initial value of I = 1 and R = 0 to represent the first COVID-19 case of Nepal at 23^rd^ January 2020. S was taken as 29 million, which is the projected population of Nepal for April 2020 [12]. Two SIR models were fitted, first with basic reproduction number (Ro) of 2.5 (global average of COVID-19) and 1.0 (to make COVID-endemic) in Nepal. To solve differential equations (i), (ii) and (iii), we used the Runge-Kutta method available in the “deSolve” package [13] in R software version 3.5.2. We also fitted SIR model for the official COVID-19 cases between 23^rd^ January and 30^th^ April as well as between 22^nd^ March and 30^th^ April 2020 and computed Ro for these periods using Limited Memory-Broyden-Fletcher-Goldfarb-Shanno (LM-BFGS) optimization algorithm in the R software. Since second COVID-19 case was reported on 23^rd^ March 2020, we also used period between 22^nd^ March and 30^th^ April 2020 to check the effectiveness of the lockdown started from 24^th^ March 2020 in Nepal. We created the change in the infected compartment for Nepal using these Ro values and derived the time-based effect of COVID-19 cases for Nepal.

Further the aggregate macroeconomic policy responses adopted by Nepal [14], tourism related employment data [15] and possible scenarios on tourism [16] was reviewed for assessing the impact of COVID-19 created disaster on economics

## Results

### Trend of the cases

From 21^st^ January 2020 till 30^th^ of April 2020, a total of 57 COVID-19 cases have been confirmed in Nepal. The increase in case was slow with only ten cases recorded between 21^st^ March and 10^th^ April, 2020, but due to sudden sharp rise in new cases in some days, the trend of daily new cases was found erratic and inconsistent (S1 File). The growth curve of the total cases in Nepal resembles a logistic curve where the total number of cases and the curve for active COVID-19 persons have also risen in similar manner but a dip in active cases have been observed lately (S2 File). The number of recoveries has also risen significantly in the recent past two weeks which is a positive sign.

In Nepal, male with age between 21- 30 years are found more infected. Highest number of infected male populations may be related to the cases dominated from the male who lived in Bhulke, Triyuga Municipality, Udayapur district and the person who came in contact with them. Majority of infected belong to the people who entered Nepal from India while some of the infected had a history of travel from other countries such as China, France, UAE, Belgium, UK and Saudi Arabia [7].

From the trend graph of RT-PCR tests (S3 File) the percent of cases have rapidly decreased as the number of tests increased in the month of February and has remained consistently low between 0.2 to 0.5 % thereafter.

### Doubling time of COVID-19 cases in weekly data analysis

Examination of doubling time of the COVID-19 cases in Nepal based upon weekly data analysis showed that there has been an increase in doubling time steadily starting around third week of March till second week of April from 3.5 to 14 days with a long span of time for doubling duration initially from one reported case to two. Thereafter, it dropped sharply followed by unexpected sudden rise in the cases during the third week of April. After that, it has again increased to around 11 days showing a slowing down of trend of the infected persons. The overall picture shows that there have been ups and downs in the doubling time of the COVID-19 cases in Nepal and is currently around 11 days (Fig 1)

**Fig 1.**
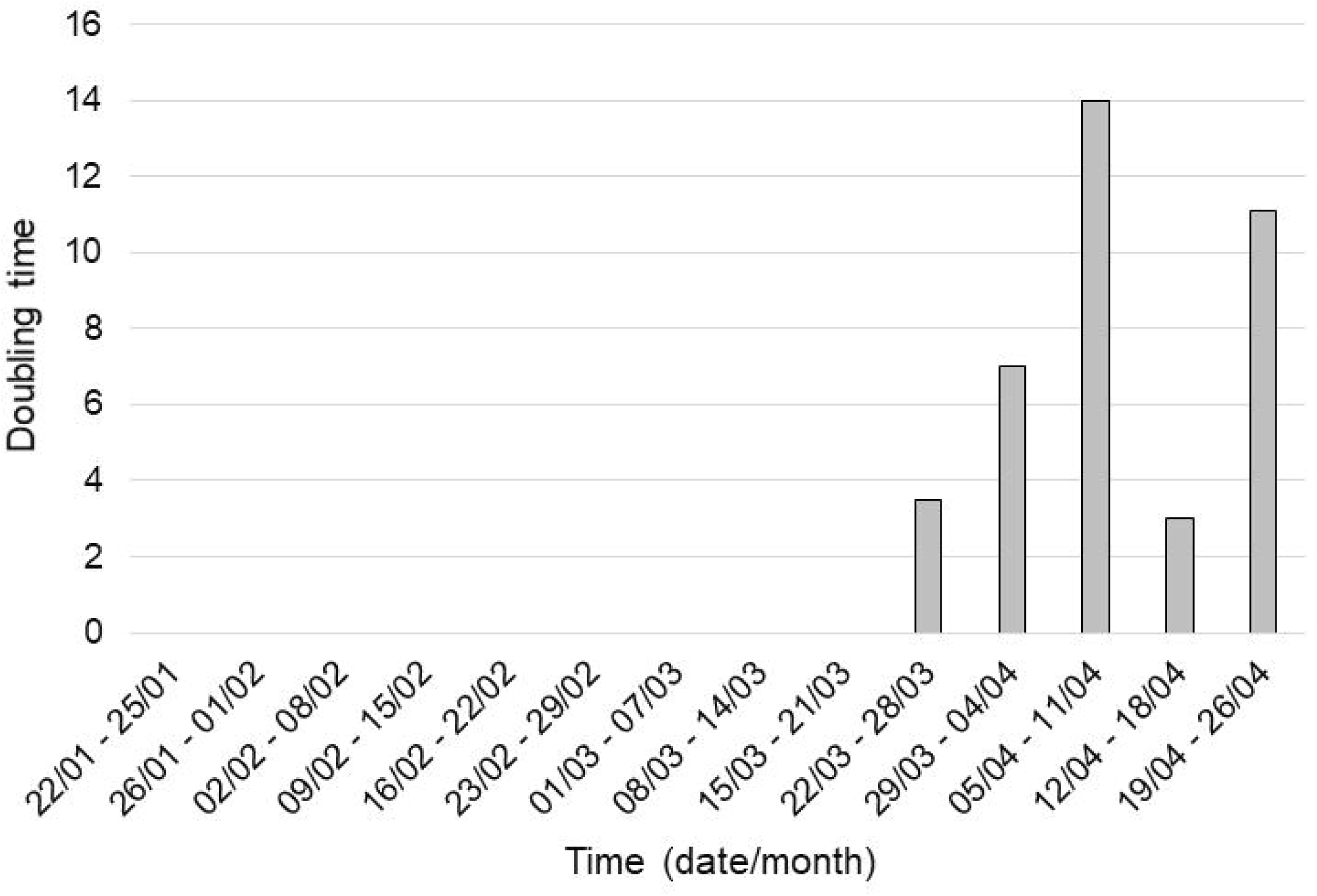
Doubling time of COVID-19 cases in Nepal (Based upon weekly data analysis from 22^nd^ January 2020 to end of April 2020

### Statistical Modelling of COVID-19 Cases of Nepal

The best ETS state-space model was found to be ETS (M, A, N) for the data i.e. exponential smoothing with **M**ultiplicative **E**rror, **A**dditive **T**rend and **N**o **S**easonality model. This is equivalent to Holt’s linear additive model (Hyndman and Athanasopolus 2018).

**Fig 2.**
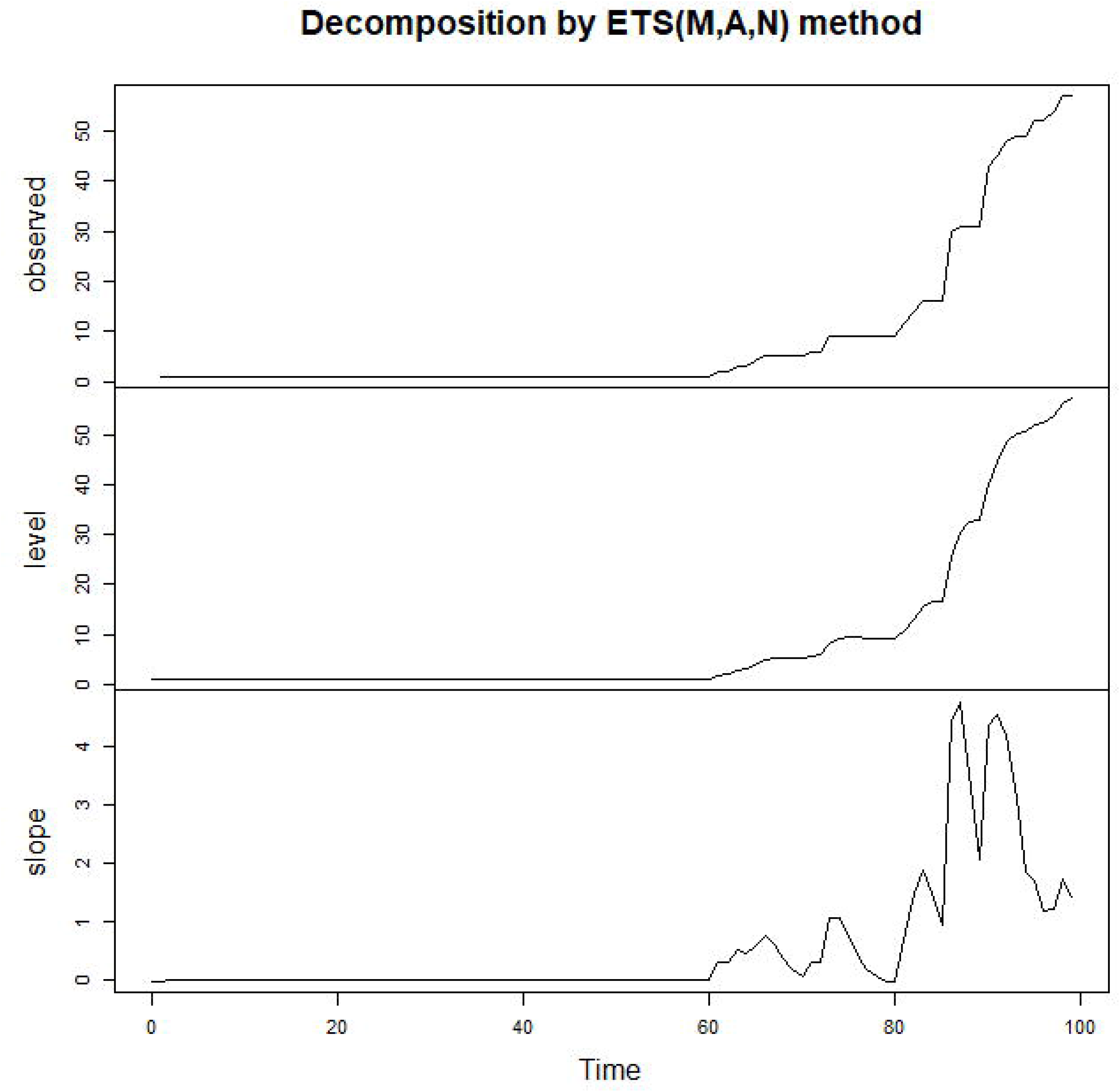
ETS decomposition of cumulative COVID-19 cases of Nepal (23^rd^ Jan. – 17^th^ April 2020)

Forcased cumulative COVID-19 cases using ETS (M,A,N) model revealed that COVID-19 cases were increasing at the rate of 2 cases per day in Nepal (Table 1). The 95% confidence interval of the forecasts were very wide, which means that starndard errors are high and precision of the estimation is low. Thus, forecasted values may not coincide with the official reported values.

**Table 1.**
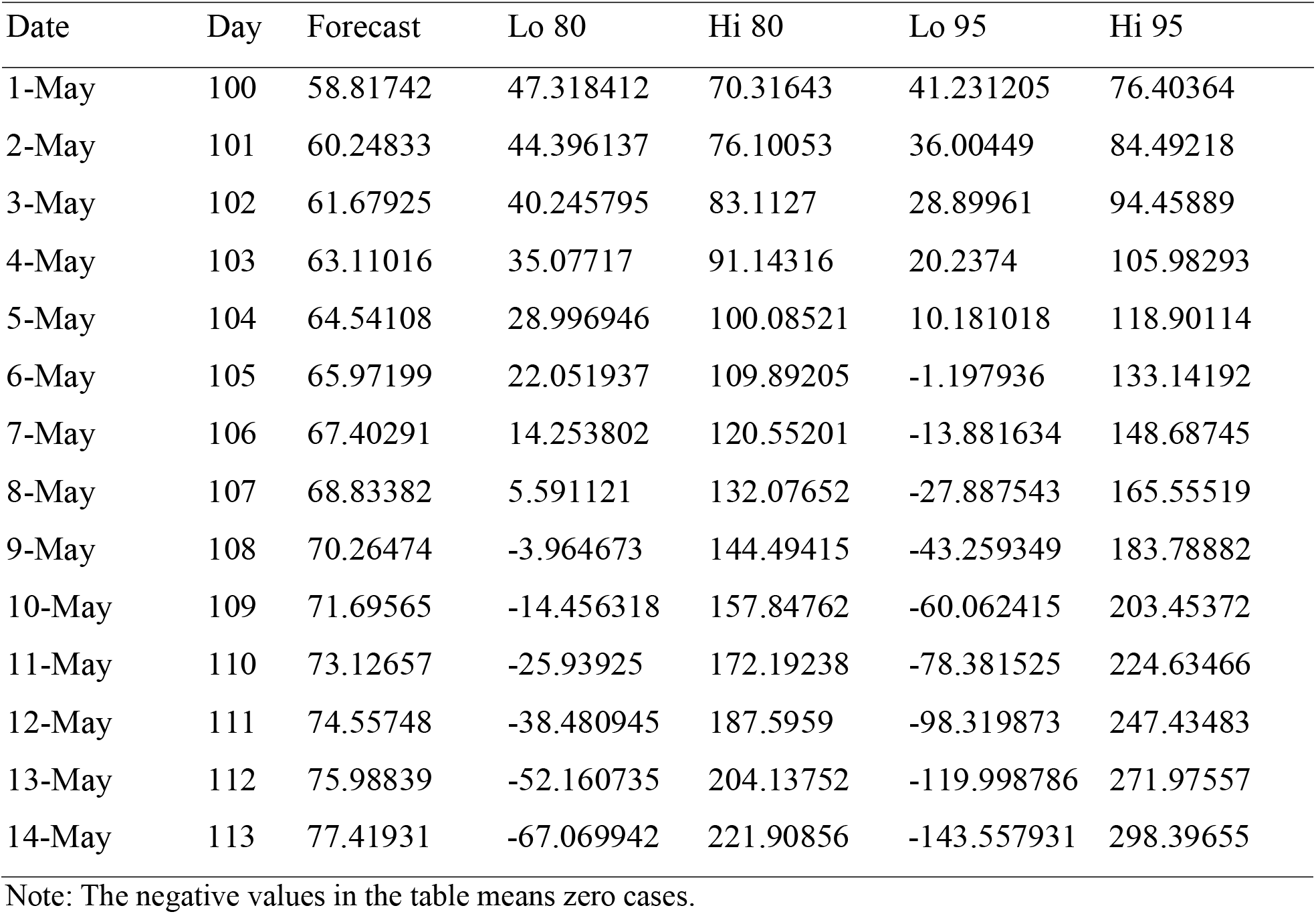
Forecasted cumulative COVID-19 cases using ETS (M,A,N) model, Nepal.

The cumulative COVID cases in Nepal revealed the increasing trend of COVID-19 cases with 205 wide variation (Fig 3).

**Fig 3.**
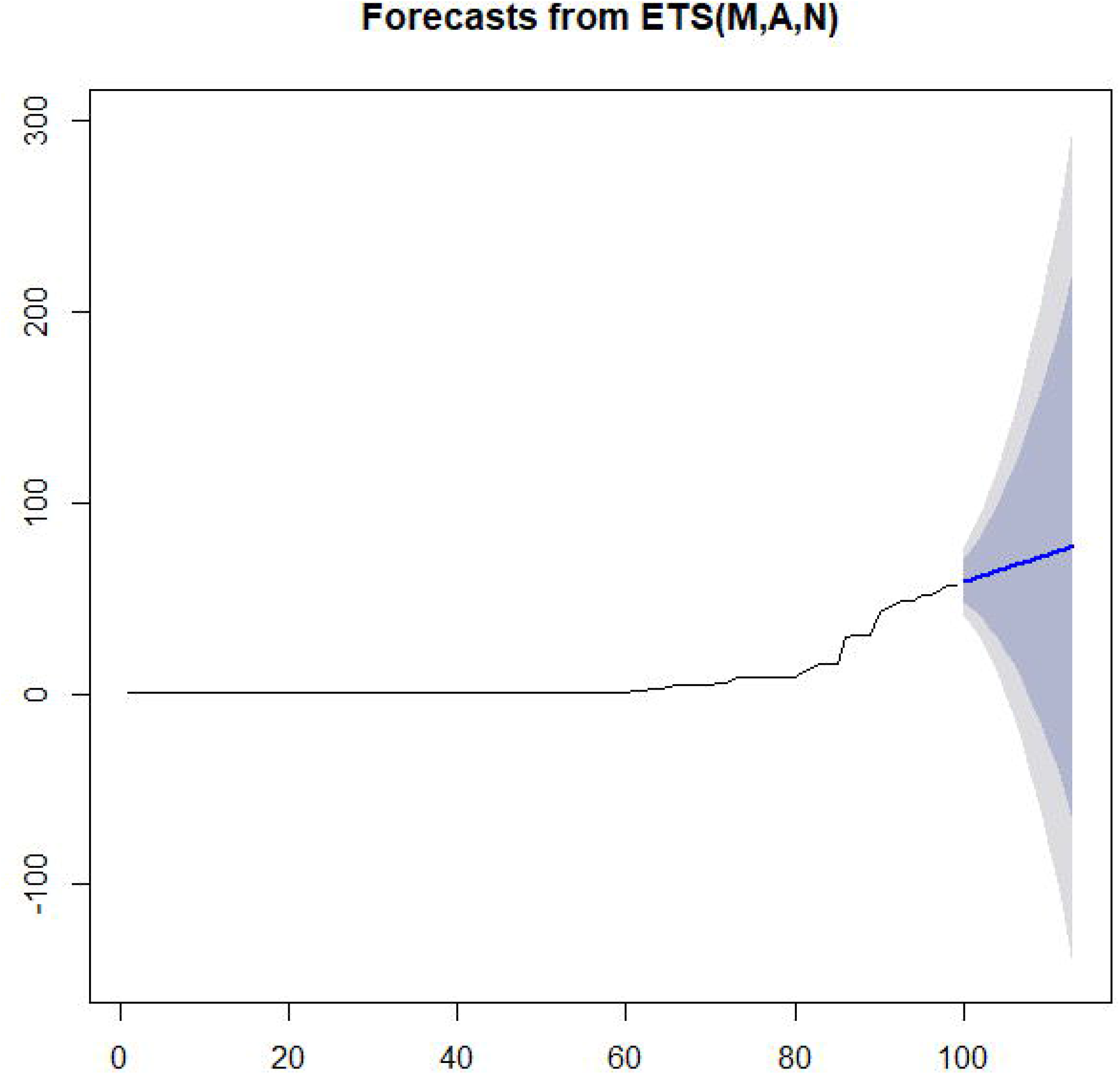
Cumulative COVID-19 cases of Nepal: Jan23 – 1 May 2020 using ETS (M,A,N) model showing increasing trend

The best ARIMA model for this 99-day data of Nepal is ARIMA (3,2,1). This means it required two times difference of the series first to make the data stationary followed by third order autoregressive filter and first order moving average filter to correct the autocorrelation and forecasting error in the data. Forecasted cumulative COVID-19 cases using ARIMA (3,2,1) model shows the forecasts obtained from this model for the next 14-day for Nepal (Table 2).

**Table 2.**
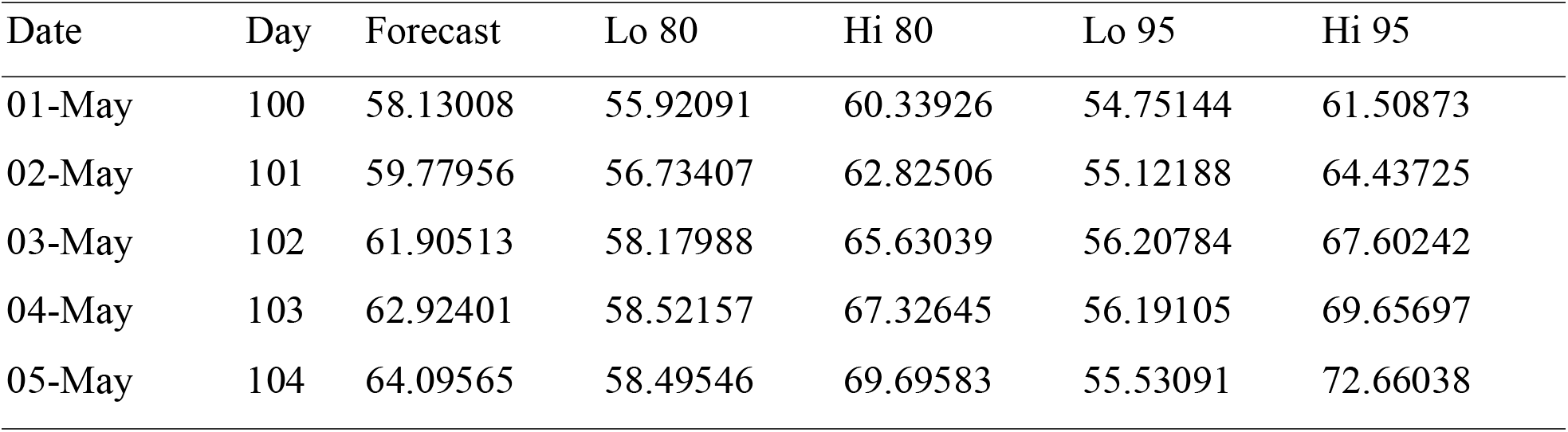
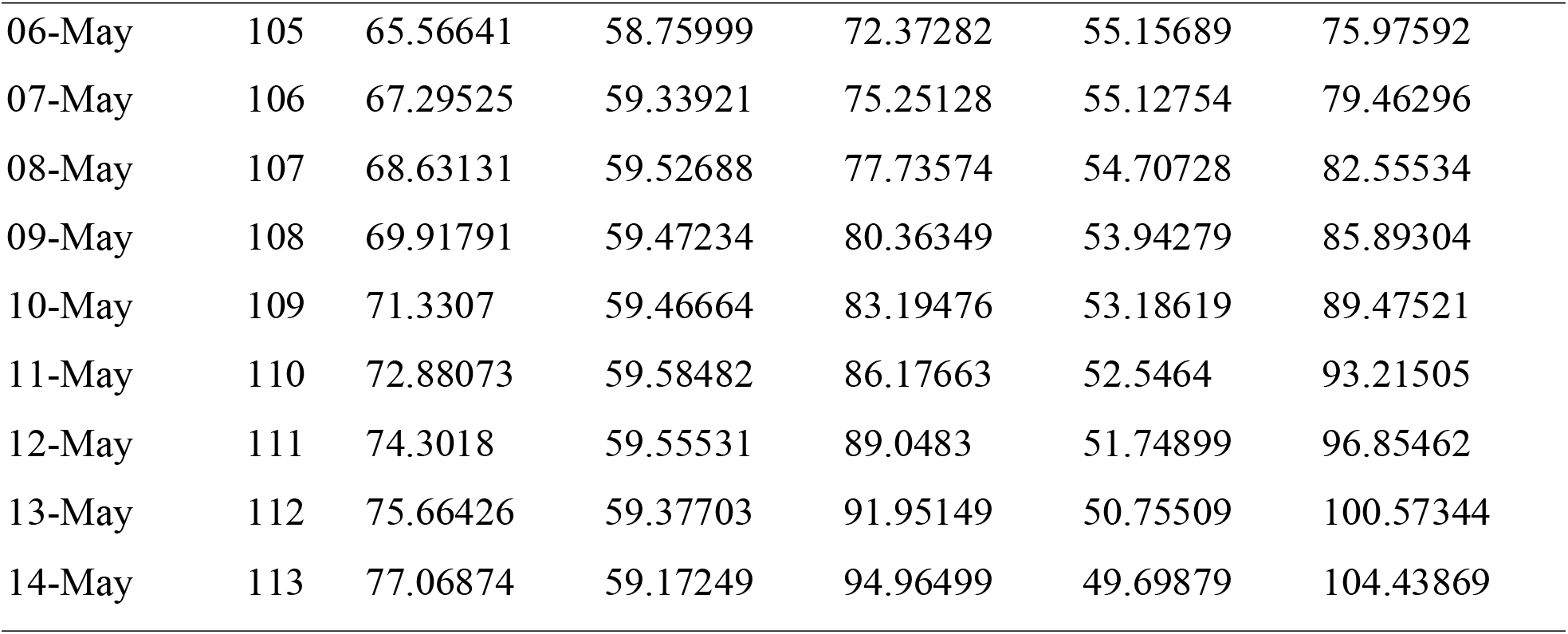
Forecasted cumulative COVID-19 cases using ARIMA (3,2,1) model, Nepal.

COVID-19 cases increasing at 2 cases per day in Nepal (Table 2). The 95% confidence intervals were found realistic. However, the forecasts may not coincide with the official reported cases.

Forecast from ARIMA (3,2,1) model were lower than the official COVID-19 cases of Nepal between 1^st^ and 14^th^ May 2020 (Fig 4). This means we need other approaches for modelling COVID-19 cases in Nepal.

**Figure 4.**
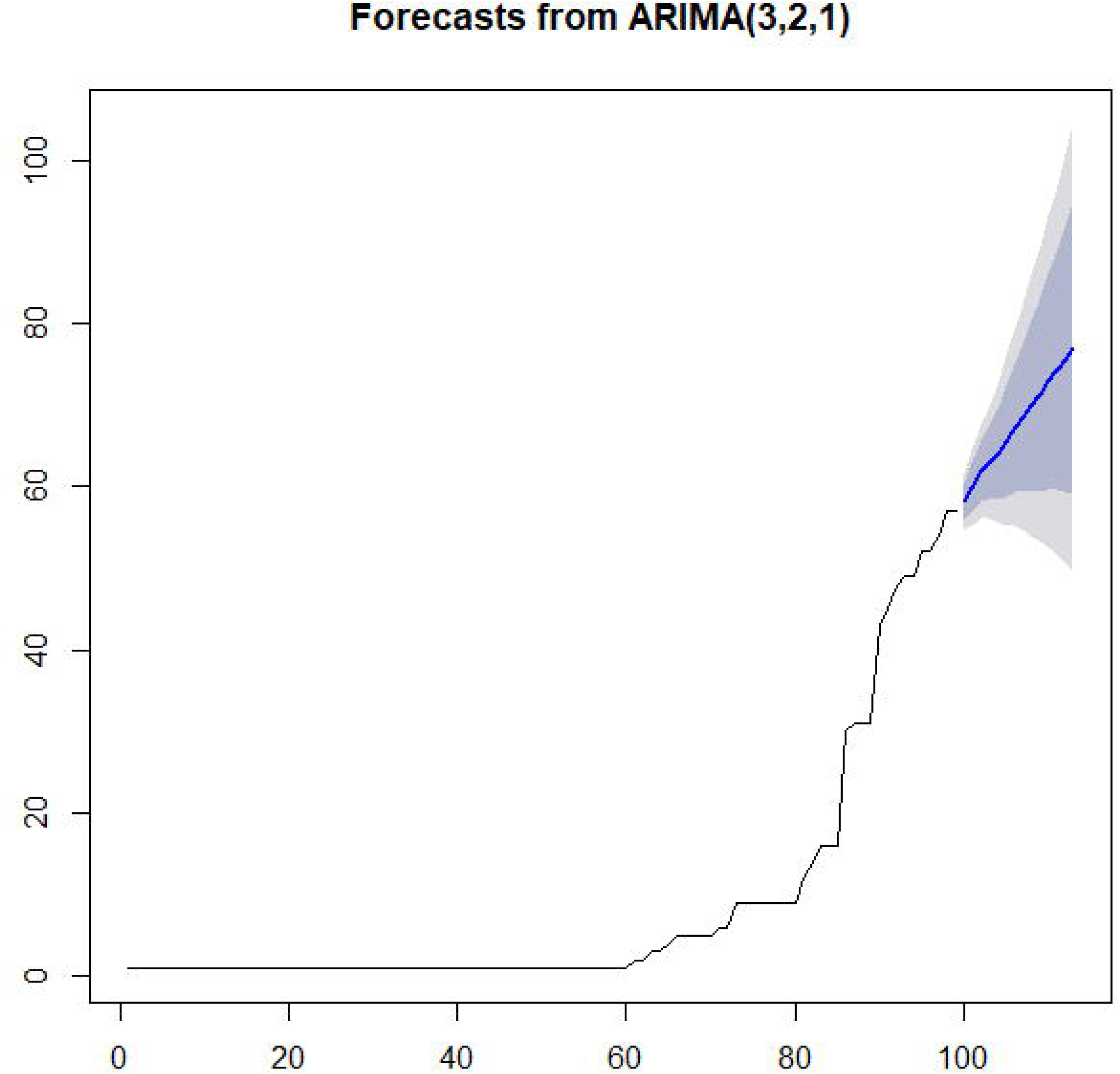
Cumulative COVID-19 cases of Nepal: 23^rd^ Jan. – 1^st^ May 2020 using ARIMA (3,2,1) model showing increasing trend

## Mathematical Modelling of COVID-19 cases of Nepal

### Scenario 1

Beta = 1/2 = 0.5 per person; Beta = 0.5/29,000,000 = 1.72 × 10^−8^ for Nepal and Gamma = 1/5 = 0.2 (reciprocal of average incubation period of COVID-19 cases) (Fig 5). This gave basic reproduction number (Ro) as 0.5/0.2 = 2.5, which is the average Ro around the world [17].

**Figure 5.**
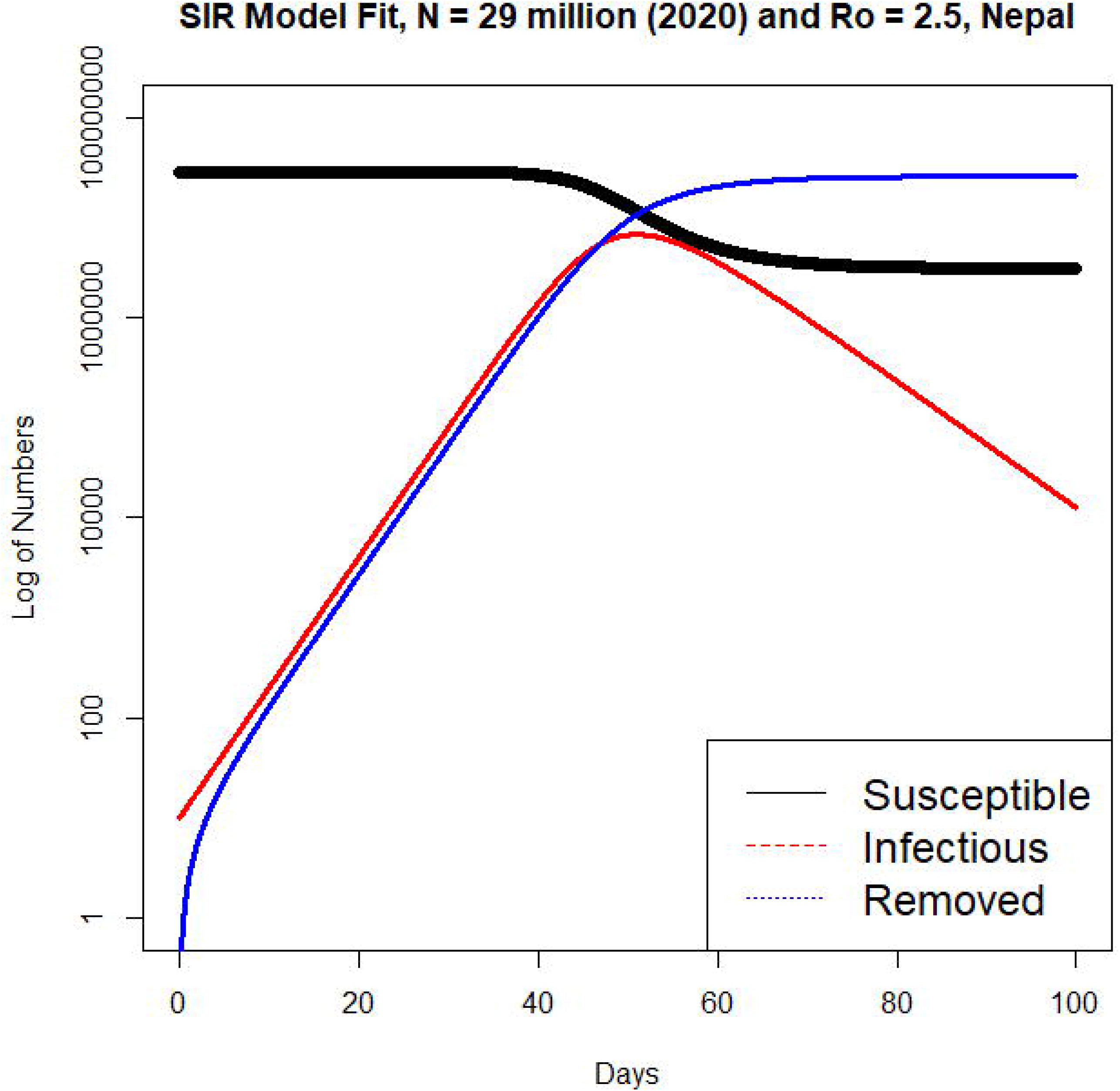
SIR model with β = 0.5 and γ = 0.2 for N = 29,000,000 of Nepal which gave basic reproduction number Ro = 2.5

This SIR model shows that 6770893 persons will be infected around 60 days whereas 38,560 persons will be infected at 100 day since the first case appeared in Nepal.

### Scenario 2

Beta = 1/5 = 0.2 per person; Beta = 0.2/29,000,000 = 6.689 × 10^−9^ for Nepal and Gamma = 1/5 = 0.2 (reciprocal of average incubation period of COVID-19 cases) (Fig 6). This gives basic reproduction number (Ro) as 0.2/0.2 = 1.

**Fig 6.**
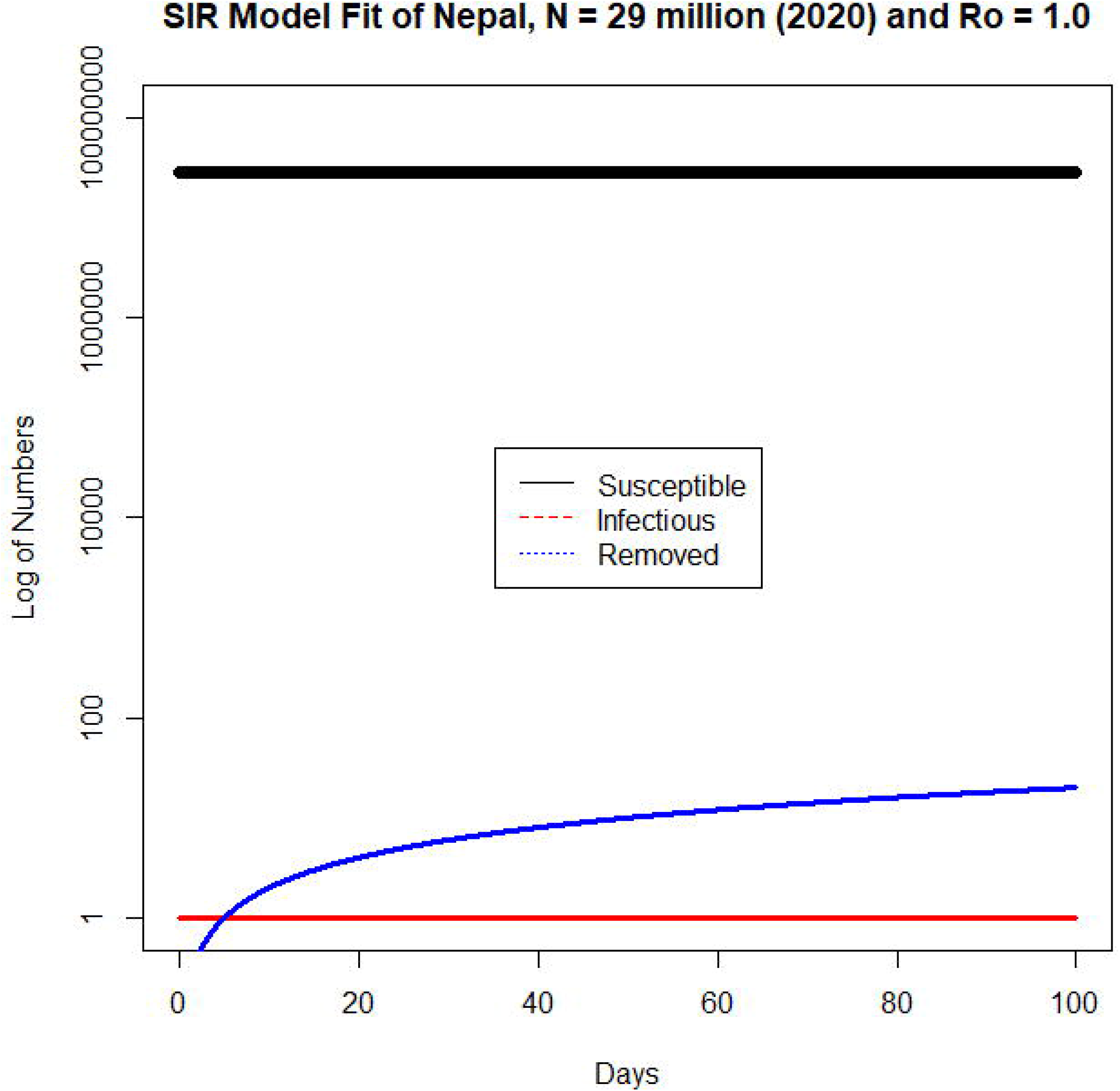
SIR model with β = 0.5 and γ = 0.5 for N = 29,000,000 of Nepal giving basic reproduction number Ro = 1

This SIR model shows that only 1 persons will be infected each day as the basic reproduction number is 1.

### Scenario 3

We used estimated β = 0.5197019 and γ = 0.4802873 obtained for official cases between 23^rd^ January and 30^th^ April 2020 using LM-BFGS optimization in R software giving Ro = β/γ = 1.08. This SIR model revealed that there will be maximum of 532,627 cases if it continues with this Ro. (Fig 7).

**Fig 7.**
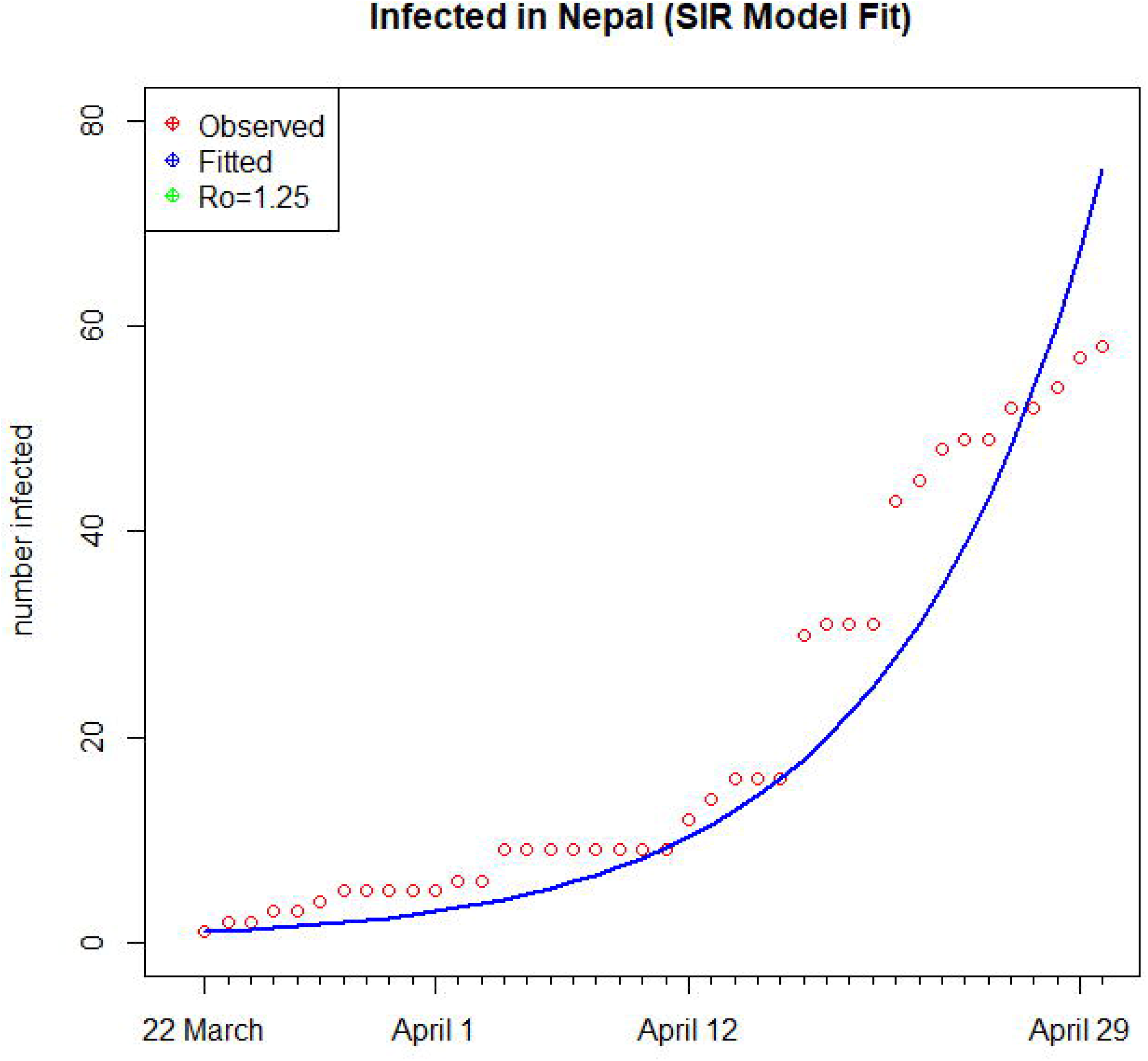
SIR model with official cumulative COVID-19 cases of Nepal if Ro = 1.25

Estimated β and γ obtained using LM-BFGS optimization in R software for official cases reported between 22^nd^ March and 30^th^ April 2020 were 0.5553948 and 0.4446052 respectively giving Ro = beta/gamma = 1.25. This SIR model revealed that there will be maximum of 6197014 if it continues with this Ro.

When Ro = 1 was considered for the official data on 30^th^ April 2020 then maximum cases will be 58.5462 and COVID-19 epidemic will be over by June 2020 at this Ro in Nepal (Fig 8).

**Fig 8.**
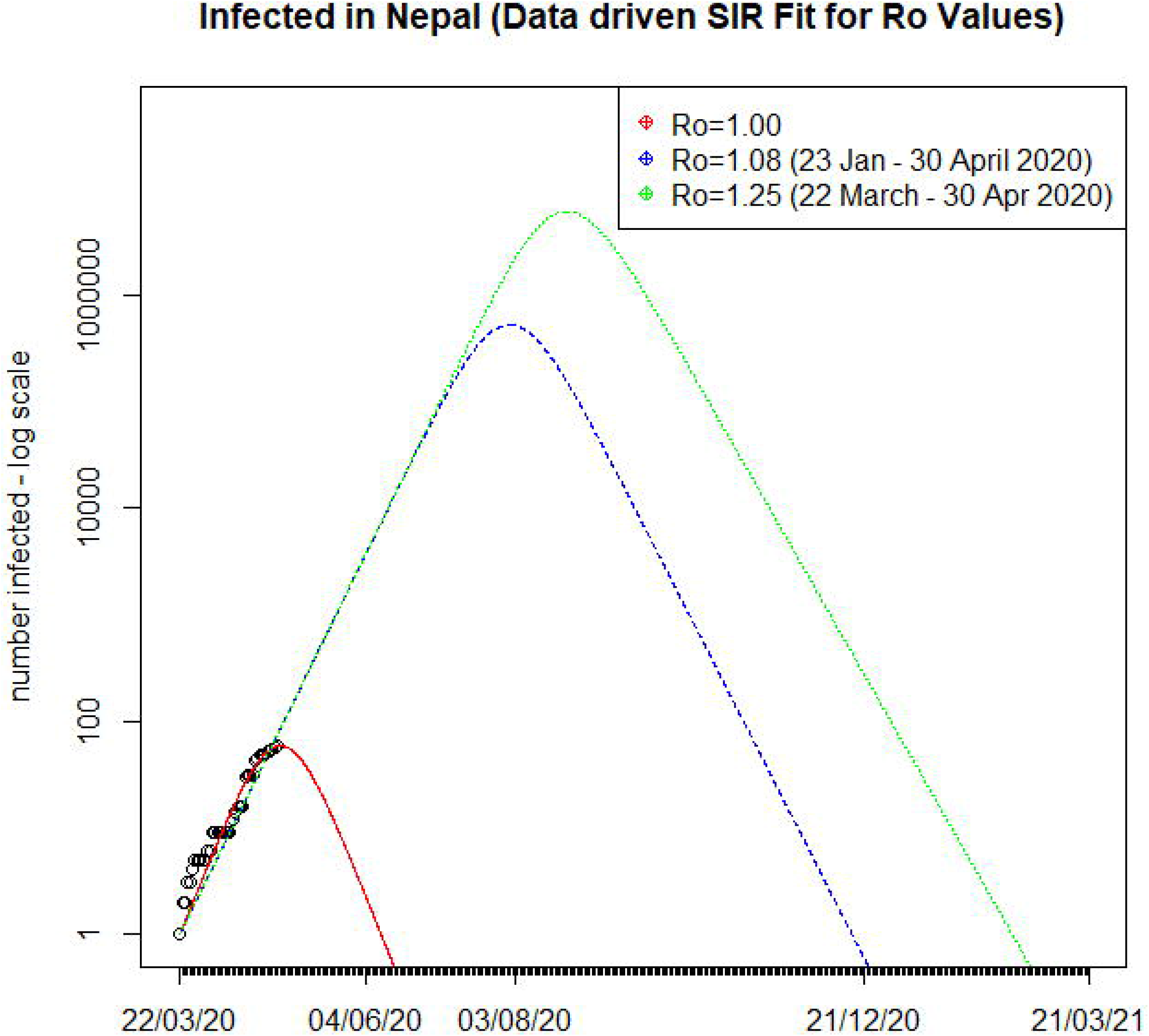
Data driven SIR model fit with three Ro values (1, 1.08 and 1.25) for Nepal

SIR model based on official data revealed that COVID-19 cases will increase in Nepal with Ro of 1.08 and the epidemic will be over only by December 2020 with peak of cases on August 2020 for this Ro. On the other hand, COVID-19 epidemic will only be over around one year time if Ro continues as 1.25.

## Impact

Tourism associated employment loss and its impact on livelihood is an external sector shock in the Nepalese economy owing to COVID-19. Among the top 10 international tourist arrival in Nepal: India, Germany, France, UK, and USA have profoundly affected economies due to COVID-19 while Australia, South Korea, China and Thailand are already on the verge of flattening the curve [18]. It indicates the decaying demand for international tourism in Nepal. The “International-Tourism-Arrival elasticities to Employment” in the case of Nepalese tourism are 0.54 for direct employment and 0.57 for the total-employment that includes indirect employment also [19]. If the restrictions on global tourism remain till end of July, international tourism arrival in Nepal will shrink by 58%. As the elasticity coefficient is 0.54, the direct tourism-related jobs will reduce by 31%. It means the tourism sector will directly result in loss of 156,000 jobs. If the COVID-19 related restriction be extended till December, international tourism will shrink by 78% and the direct tourism-related employment will reduce by 42%, which totals 210,000 jobs loss. Similarly, out of 700,000 total tourism-related jobs, 231,000 to 311,000 jobs will disapper for several months which is a scary situation for a developing economy like Nepal.

## Discussion

The analysis of COVID-19 cases in Nepal provides an insight into situation till implementation of first phase of lockdown and prediction of expected scenario thereafter. The data of the cases were based on the results of test confirmed by RT-PCR. From the trend graph of cases as percent of RT-PCR tests the percent of cases have rapidly decreased as the number of tests increased in the month of February and has remained more or less constant between 0.2 to 0.5 % thereafter. This is an indication of absence of significant rise in COVID-19 cases despite increase in the tests substantially in the months of March-April in Nepal, although the number of tests may still not be sufficient to cover large proportion of population in Nepal. The overall growth of COVID-19 cases in Nepal is not high, which can be taken positively and credited to the lockdown implemented by Nepal Government and awareness of the diseases through media along with the preventive measures. Doubling time of COVID-19 cases along with incubation period of the coronavirus are important variables in the spread of the epidemic. The incubation period of the virus has been reported with certain amount of variability (2–14 days) with the median average hovering around 5 days with possible outliers up to 27 days [20].

The overall picture shows that there has been ups and downs in the doubling time of the COVID-19 cases in Nepal and is currently around 11 days (Fig 1) which is just above 10 days doubling time of COVID-19 cases in India as reported by different news channels of India though the total cases in Nepal are still negligible compared to that of India. Bhandary (2020) has stated that with implementation of lockdown the doubling time increased from 5 days to 15 days [21].

Statistical modelling showed that COVID-19 cases may continue to increase exponentially in Nepal. Forecasts from best ARIMA model (MAPE = 4.18) are found to be more precise than the ETS exponential smoothing model (MAPE = 4.55). Forecasts from best ARIMA model will be a valid estimate if clusters with more than 10 cases will be found there in the future too. This has already happened once between 18^th^ April and 1^st^ May 2020 in Nepal and thus requires particular attention of the concerned administrators and policy makers. However, it will not be valid if the cases will be increased in higher rates in the country. ETS and ARIMA models produced estimated values of COVID-19 cases for Nepal based upon the currently available official data. Despite the possibility of questions on whether sufficient tests have been conducted for such modelling, COVID-19 cases in Nepal has not risen sharply in high numbers and the total cases in Nepal are still below 100 but expected to rise in coming days.

Compartmental SIR model on the other hand reveals that nearly seven million Nepali population might be infected with average global basic reproduction number of 2.5 for COVID-19 cases and maximum cases would have reached within 60 days of the first case, which fortunately did not happen in the country. The total number of cases would have been only 1 if the basic reproduction number remained at one in Nepal, which also did not happen as 30 cases were reported by 17^th^ April 2020 and 58 in 30^th^ April 2020.

Basic reproduction number of Nepal was computed as 1.08 for 23^rd^ Jan – 30^th^ April 2020 period, which means lockdown was working well to maintain low level of basic reproduction number in the country. Further, it also revealed that COVID-19 will only be over by December 2020 with peak cases in August 2020 with Ro of 1.08. On the other hand, basic reproduction number of 1.25 was computed for total cases reported from 22^nd^ March to 30^th^ April 2020 period, which means that COVID-19 did not increase exponentially but it will remain for at least a year in the country. Thus, these results clearly suggests to continue the lockdown in the hotspots (heavily affected areas) and slowly open the lockdown on least affected areas with social/physical distancing and personal hygiene maintenance in Nepal.

This analysis we performed have been restricted to the lockdown implemented from 24^th^ March 2020 till the end of April 2020 and not a final assessment yet considering that the spread of disease could still be at the early stage in Nepal and calls for re-assessments periodically in future as well till the pandemic phases out.

Considering the containment of outbreak lockdown is necesary, it will have huge impact in the country’s economy and the possible scenario of easing international travel has been contemplated. Based on the [22], the best-case scenario assumes a gradual opening of international borders and easing of travel restrictions in early July. It will witness reduction in global tourism arrival likely by 58 per cent. The second scenario assuming the international tourism arrival shrinkage by 70 per cent points is due to the restriction in situations goes until early September. Relatively uncomfortable third scenario explains the easing of the restriction sustains till December with arrival number reduction by 78 per cent. Considering these three optimistic scenarios, we relate the international tourism arrival and employment plausibility in Nepal.

The tourism sector faced a sharp downswing since the COVID-19 outbreak and with uncertainty continued to dominate. The tourism sector will loose about US$ 910 billion to US$ 1.2 trillion in export revenues from tourists. Most importantly, about 100 to 120 million direct tourism related jobs are at stake.

The tourism sector contributes about 2% of the GDP and some 15–20% of the foreign reserve. Furthermore, this sector is generating a significant volume of employment in Nepal. In Nepal, there are about 500,000 direct and 700,000 total jobs in the tourism sector. Nepalese tourism sector is one of the livelihood options to many marginal people. When the lockdown struk the tourism sector, the livelihood issues arose that informal or unorganized job holders will experience hardest hit.

The unprecedented lockdown also makes domestic tourism very unlikely, which means that slowdown of global tourism is going to be the decisive factor influencing employment in the tourism sector in Nepal.

## Conclusions

The data based trends for COVID-19 epidemic in Nepal have been analyzed followed by assessment of the outbreak impacts in comparison with existing models. The outcomes of the research can be summarized as follows:

1. The total COVID-19 cases are increasing by following the logistic curve. However, there has been a variation in the doubling time during the analysis period, which can be attributed to the effect of lockdown imposed by the Government. The forecasts from best ARIMA model may provide more precise and valid estimates if clusters with more than 10 cases are found in future.
2. As in compartmental SIR model, nearly 7 million people might get infected. With an average global basic reproduction number of 2.5 for COVID-19 cases, maximum cases would have reached within 65 days, which fortunately did not happen in the country. The basic reproduction number in Nepal being maintained at low level of 1.08 for the period between 23^rd^ January and 30^th^ April, 2020 is an indication of effectiveness of the lockdown in containing the COVID-19 spread.
3. Further, the model has suggested that the COVID-19 will only be over by December 2020 with the peak cases in August 2020. On the other hand, basic reproduction number of 1.25 was computed for total cases reported for 22^nd^ March – 30^th^ April 2020 period, which implies that COVID-19 may persist at least for a year in the country. Thus, the results so far obtained clearly recommend continuing the lockdown in the hotspots (heavily affected areas) and gradually easing the lockdown in the least affected areas with strict social/physical distancing and personal hygiene maintenance.
4. Since the COVID-19 is expected to destroy the Nepalese economy on many fronts a robust impact study on economic impact of COVID-19 in Nepalese economy is suggested.

## Data Availability

The data will be available when requested for

## Acknowledgements

We are thankful to Prof Dr. Dinesh Raj Bhuju, Academician, Nepal Academy of Science and Technology for his encourgament. We acknowledge Ministry of Health and Population, Nepal Government for the data on COVID-19.

## Supporting information

**S1 Fig. Trend of daily new cases and recovered cases of COVID-19 in Nepal**

**S2 Fig. Trend of active, recovered and total number of COVD-19 cases with time**

**S3 Fig. Number of cases as percentage of total PCR tests conducted on the samples from COVID-19 suspected**

## Notes

### Competing Interest Statement

The authors have declared no competing interest.

### Clinical Trial

This study does not have any clinical trial

### Funding Statement

No funding has been received for this work

### Author Declarations

No human sample or subjects were involved so ethical approval from IRB is not required for this study

